# *CDH13* is associated with cellular viability after exposure to ionizing radiation using genome-wide screening

**DOI:** 10.64898/2026.06.12.26355511

**Authors:** Hannah-Lena Schmidt, Olena Ohlei, Sarah Herwest, Bastian Salewsky, Lars Bertram, Ilja Demuth

## Abstract

**Background:** It is well known that genetic variants contribute to cellular sensitivity to chemotherapeutic agents and ionizing radiation (IR). The aim of this study was to identify single nucleotide polymorphisms (SNPs) and genes associated with the spectrum of normal cellular sensitivity of lymphoblastoid cell lines (LCLs) towards ionizing radiation and mitomycin C (MMC).

**Methods:** In a first step, we determined the viability of LCLs established from male participants of the Berlin Aging Study II (BASE-II) aged ≥62 years following treatments with increasing doses of IR (n=137 cell lines) or MMC (n=140 cell lines) using the alamarBlue assay. Results from intra-experimental triplicates and three independent experiments for each cell line and treatment were used to calculate the area under the curves (AUCs) representing the specific sensitivity to IR and MMC of each LCL. The data from these experiments were subsequently used as outcomes in genome-wide association studies (GWASs). In addition, we calculated polygenic risk scores (PGS) from UK Biobank GWAS results for four cancer-related phenotypes and assessed the extent to which the variance in the IR and MMC sensitivity is explained by these PGS.

**Results:** The GWAS analyses revealed one variant, rs74728080, located in *CDH13* on chromosome 16, to show genome-wide significant (p < 5 × 10-8, ß = 2.81) association with cellular viability after treatment with IR. In the GWAS on MMC sensitivity the most interesting signal was elicited by SNP rs113978558 in an intron of the *PLD5* gene on chromosome 1 (p = 9.232 × 10-8; ß = 1.44). Several other SNPs with statistically suggestive (i.e., p < 1 × 10-5) evidence of association with IR or MMC sensitivity were identified. PGSs calculations from GWAS of four cancer-related traits in UKB explained ∼5% and ∼3% of phenotypic variance in IR- and MMC-induced cell viability, respectively.

**Conclusion:** The genome-wide significant association of rs74728080 with IR sensitivity and the location of this variant in *CDH13* is interesting and functionally highly plausible given its known involvement in oxidative-stress response and function as tumor suppressor. Taken together, our novel data suggest that *CDH13* may be genuinely involved in regulating cellular IR sensitivity.

## Introduction

Radiotherapy (RT) is used for cancer treatment in a large proportion of patients at some point during the course of their disease [1, 2]. To a lesser extent and in a smaller spectrum of cancer entities, mitomycin C (MMC) is an important component among available anti-tumor therapeutics as well [3]. While therapeutic options, e.g., in terms of hit accuracy at the target tumor have steadily improved (reviewed for radiotherapy in [4]), short and long-term side effects still remain a complex and clinically highly relevant problem with limited possibilities to be predicted before starting the therapy. It has been widely accepted that normal tissue radiosensitivity is, at least partly, determined by genetics [5–10]. This idea was initially supported by findings on rare monogenetic diseases, where the basic defect was found in genes involved in DNA damage signaling or DNA repair, such as Ataxia telangiectasia [6, 8], Nijmegen breakage syndrome [5, 9] and Fanconi anemia [7, 10]. While these diseases are typically the consequence of two mutated alleles, cells from people heterozygous for DNA sequence variants in this group of genes were shown to display increased sensitivity to ionizing radiation (IR), radiomimetics and/or DNA-interstrand crosslinking agents like MMC (reviewed for IR in, e.g., [11–15]). Consequently, multiple candidate gene studies assessing the role of single nucleotide polymorphisms (SNPs) in known DNA damage response genes have been reported (e.g., [16–18]). Overall, however, these studies led to inconsistent results and some suffered from methodological shortcomings [19]. Improvements in high-throughput DNA genotyping techniques now allow to search for novel disease genes through genome-wide association studies (GWAS). This strategy was also adopted by the field investigating adverse outcomes related to radiotherapy and chemotherapy as well as to cellular sensitivity in response to IR and substances used in cancer treatment [e.g. 20-23]. These GWAS were conducted for a variety of cancer sites and were aiming to identify loci associated with therapy-induced toxicities. For instance, Kerns et al. reported a meta-analysis including 3,871 prostate cancer patients from six GWAS datasets who received RT. The authors identified four independent genome-wide significant loci associated with various radiotoxicities (decreased urinary stream, rectal bleeding and/or hematuria) [20]. Similarly, a GWAS on 972 multiple myeloma patients who received high-dose chemotherapy including alkylants, which have a similar mode of action as MMC, and autologous stem cell transplantation revealed 11 SNPs showing strong association with severe or life-threatening oral mucositis [21]. Inclusion of these SNPs increased the sensitivity in predicting oral mucositis by nearly 10% beyond known risk factors [21].

In a few studies, the GWAS approach was used to identify SNPs associated with the spectrum of normal cellular sensitivity to chemotherapeutic agents and IR in Epstein-Barr virus-transformed T lymphocytes, e.g., lymphoblastoid cell lines (LCLs). In a first step, these studies evaluated cellular viability following treatment with the mutagen of interest, typically assessed as the area under the curve (AUC) or the maximal inhibitory concentration (IC50) following treatment with increasing doses. Subsequently, these outcomes – which quantify the cellular mutagen response – were assessed by GWAS. For example, the study by Niu et al. [22] identified 27 loci associated (at p < 10^-4^) with IR sensitivity assessed in 277 LCLs. The authors additionally showed that 14 of these loci were associated with 39 genes whose mRNA expression was also associated with IR sensitivity. Furthermore, the authors showed that a subset of these genes was involved in IR sensitivity following siRNA knockdown experiments in cancer cell lines [22]. In a similar approach, Mulford and colleagues [23] used the GWAS approach with data quantifying the sensitivity (IC50 or AUC) to eight different chemotherapeutic drugs assessed in 164 to 432 LCLs from three 1000 Genomes Project ancestries. These analyses revealed four independent loci associated (at p < 5 × 10^-8^) with cellular sensitivity to daunorubicin, carboplatin, etoposide, or cisplatin. Interestingly, none of these associations were observed with LCLs derived from individuals of European descent. Similarly, GWASs based on cellular sensitivity to cisplatin or derivatives of cyclophosphamide revealed no associations below the genome-wide significance level [24, 25]

In summary, these earlier studies did not uniformly use stringent genome-wide significance thresholds, did not reveal associations in cell lines established from people of European ancestry, and/or did not investigate MMC sensitivity. To close this gap, we performed a GWAS on MMC sensitivity in cells derived from European individuals using established (and conservative) thresholds for determining genome-wide significance. To this end, we generated cellular sensitivity data using IR and MMC as exposures in a total of 137 and 140 LCLs, respectively. Subsequent GWAS analyses identified a variant (rs74728080) in the *CDH13* gene showing genome-wide significant association (p < 5 × 10^-8^) with cellular viability after IR treatment. *CDH13* is a compelling candidate in this context due to its protective role in oxidative stress-induced apoptosis (reviewed in [26]) and cancer development (e.g., [27–33]).

## Methods

### Cell lines and cell culture

Out of a total of 432 LCLs of participants of the Berlin Aging Study II (BASE-II) that had previously been established and were available for further examination [34], 140 LCLs from men in the older BASE-II subgroup were randomly selected for this project. BASE-II is a multidisciplinary and multi-institutional project investigating factors associated with “healthy” vs. “unhealthy” aging. The baseline cohort is represented by 2,200 adult volunteers from the Berlin metropolitan area. Of these, 1,600 participants were assigned to the “older” subgroup (aged 60-80 years), whereas the remaining 600 individuals were assigned to the “younger” subgroup (aged 20-35 years), serving as reference population. More detailed information about BASE-II can be found in previous work from our group [35–37]. All participants gave written informed consent. The Ethics Committee of the Charité – Universitätsmedizin Berlin approved the study (approval numbers EA2/029/09 and EA2/144/16). The study was conducted in accordance with the Declaration of Helsinki and was registered in the German Clinical Trials Registry as DRKS00009277.

The LCLs were cultured in a humidified incubator at 37 °C with 5% CO_2_ and 21% O_2_ (BINDER GmbH, Tuttlingen, Germany) and they were grown in RPMI 1640 medium (Gibco by Thermo Fisher Scientific Inc., Waltham, Massachusetts, USA) supplemented with 15% fetal calf serum (FCS;PAN-Biotech, Aidenbach, Bavaria, Germany) and 1% penicillin/streptomycin (Gibco by Thermo Fisher Scientific Inc., Waltham, Massachusetts, USA; PAN-Biotech, Aidenbach, Bavaria, Germany).

The 140 selected LCLs were divided into two batches of 70 LCLs. The growth inhibition experiments of batch 1 were conducted as part of earlier work [34] while the growth inhibition experiments of batch 2 were newly conducted for this study using the same experimental protocol.

### Growth inhibition – mitomycin C and ionizing radiation viability assays

In summary, a total of 140 cell lines (LCLs) established from male participants (mean age 67.9 ±3.2 years) were evaluated for their sensitivity to MMC and 137 LCLs were examined for their sensitivity to IR. For the growth inhibition experiments we followed the protocol that has been previously described for the batch 1 samples [34]. Briefly, cells were harvested in logarithmic growth, counted, and then plated in 96-well flat-bottom cell culture plates (Falcon by Corning Life Sciences, Corning, New York, USA).

For the MMC viability assays, the LCLs were plated in triplicates, resulting in two 96-well plates per experiment. The first row of each plate was filled with 100 µl of cell culture medium per well only and served as negative control. After 24 hrs of incubation, the cells were treated with increasing concentrations of MMC (0, 1, 5, 50, 200, 500 and 1000 nM; Sigma-Aldrich, St. Louis, Missouri, USA) by adding 100 µl of the applicable twofold concentrated MMC solution. Each concentration of the MMC solution was obtained by diluting MMC in cell culture medium and then establishing a dilution series. Again, only 100 µl of cell culture medium was added to each well of the first row of the plates [34].

For the IR viability assays, the cell lines were also plated in triplicates. Separate plates were used for each radiation dose, resulting in seven 96-well plates per experiment. Here again, the first row of each plate was filled with 100 µl of cell culture medium per well only. Additional 100 µl of cell culture medium were added to each well. After 24 hrs of incubation, the cells were irradiated with increasing doses of IR (0, 1, 2, 5, 7.5, 10 and 20 Gy; GSR D1, Gamma-Service Medical GmbH, Leipzig, Saxony, Germany) [34].

After another 144 hrs (6 days) of incubation at 37 °C, both, MMC and IR viability assays were stained with 20 µl (1 : 10 dilution) of alamarBlue Cell Viability Reagent (Invitrogen by Thermo Fisher Scientific Inc., Waltham, Massachusetts, USA) [34]. Resazurin is the active component of alamarBlue. This non-toxic, cell-permeable dye is blue and non-fluorescent. In metabolically active cells, resazurin is reduced to resorufin, which is red in color and highly fluorescent [38].

After staining, the treated cells were once again incubated at 37 °C for another 24 hrs. Subsequently, the absorption of light at 570 nm and 600 nm was measured using an absorbance spectrometer (Sunrise, Tecan Group Ltd., Männedorf, Switzerland) and the cellular viability was calculated following the instructions of the alamarBlue user manual. For subsequent analyses, we pooled the data from all replications for each cell line and treatment (MMC or IR). Finally, GraphPad Prism 7 (GraphPad Software Inc., San Diego, California, USA) was used to calculate the AUC of the viability curves for each cell line, with greater AUC values indicating higher cell viability after exposure to mutagens and thus lower cellular sensitivity to MMC/IR. A schematic overview of the experimental setup is shown in figure 1.

**Figure 1:**
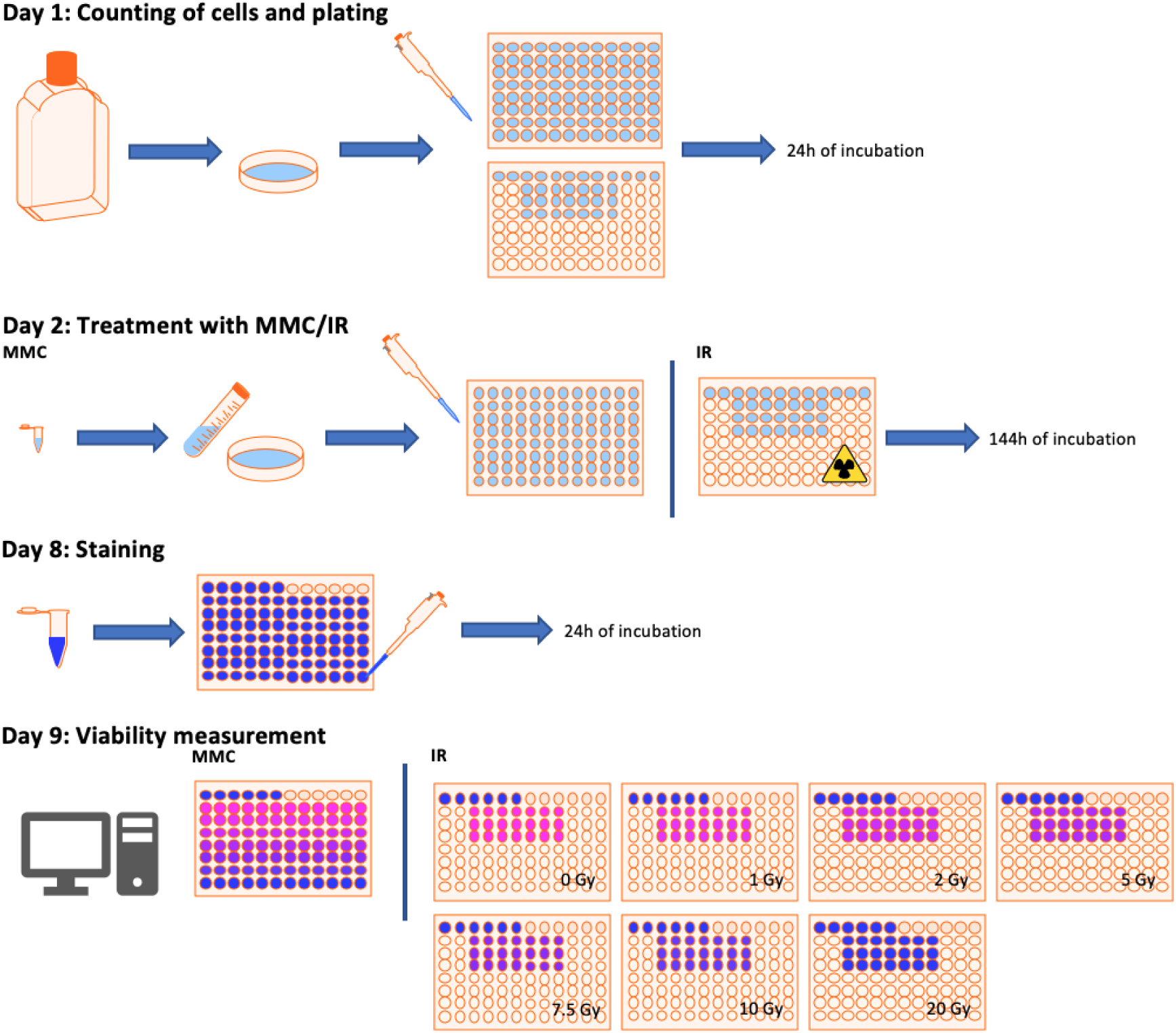
Experimental setup. After cultivation of the LCLs, the cells were counted, diluted, and seeded in triplicates on 96-well plates. After 24 hrs, the cells were treated with increasing concentrations of MMC or IR doses, respectively. One week later, the resazurin containing alamarBlue Cell Viability Reagent was added to the samples. After another 24 hrs of incubation, cell viability was measured with an absorbance spectrometer.

### Genotyping, data processing and quality control

For the GWAS analyses, DNA was extracted from whole blood samples of BASE-II participants followed by genome-wide SNP genotyping on the Affymetrix 6.0 array using procedures described previously [39]. Autosomal genotypes were then subjected to extensive quality control (QC) and imputation using an automated computational workflow [40, 41]. Imputation of untyped variants was performed on the QC’ed data using MiniMac3 software [42] using the Haplotype Reference Consortium (HRC; v1.1 [EGAD00001002729 including 39,131,578 SNPs from ∼11K individuals]) reference panel [43]. This procedure resulted in 39,131,578 imputed SNPs in all 140 participants included in this study. Prior to statistical analyses, post-imputation QC was performed at both the SNP and individual levels excluding SNPs failing to fulfil the following criteria: location on autosomes, MiniMac3 imputation quality R^2^ ≥ 0.3, MAF ≥ 0.01, and Hardy-Weinberg equilibrium (HWE) p-values ≥ 5 × 10^-6^. Furthermore, we removed samples that were related up to the fourth degree (--king-cutoff 0.025), as well as samples with excessive heterozygosity (exceeding six standard deviations (SD) from the mean). Overall, this resulted in a dataset of 7,238,717 high-quality autosomal SNPs in 133 individuals available for downstream analyses.

DNA variants on the X-chromosome were generated using the Infinium Global Screening Array (GSA; Illumina Inc., San Diego, California, USA) on the same samples. Before imputation we performed QC for pseudoautosomal regions (PAR1 and PAR2) analogously to the variants on autosomes (see above). For the remaining non-PAR variants, we performed QC separately in females and males using a workflow previously described by our group [44]. Imputation for X-chromosome variants was performed on the Sanger Institute imputation server using the same HRC reference panel as for autosomes [43]. After imputation, we removed SNPs separately for female and male data sets (in the full BASE-II data, which also included females) with MAF < 0.01 and R^2^ < 0.7. A HWE test was performed on combined samples as recommended previously [45] and implemented in PLINK2 [46]. To account for the missing second X-chromosome in males, we coded markers as 0 vs. 2 (instead of 0 vs. 1) as recommended by Smith et al. [47]. This resulted in 203,356 X-chromosome variants available for 96 individuals in the BASE-II dataset. The smaller number of samples is explained by the overall smaller number of BASE-II samples analyzed via the GSA array.

### GWAS and statistical analyses

Raw data for each treatment batch were normalized using z-transformation. GWAS analyses were then run for each batch separately using linear regression in PLINK2 accounting for three ancestral principal components calculated from a principal component analysis (PCA) on linkage disequilibrium (LD) pruned autosomal SNP data. We did not include sex as a covariate because all cell lines used in the treatment experiments were established from male participants. GWAS results for both experimental batches for each treatment were meta-analyzed using the inverse variance weighting (“fixed effect”) model implemented in METAL [48]. SNP-based GWAS results were then subjected to post-GWAS analyses, primarily using the FUMA toolkit (v1.8.3; http://fuma.ctglab.nl/; [49]). This included the estimation of gene-based effects by aggregate effect sizes of all SNPs within specific genes based on MAGMA v1.08 [50] as implemented in FUMA. We also used FUMA to select independent genome-wide significant SNPs, to annotate SNPs to their nearest genes, and to visualize GWAS results on MMC or IR treatment statuses. Alpha levels for the SNP-based analyses were set to 5 × 10^-8^, an established threshold in the context of GWAS in European populations [51], and to 2.62 × 10^-6^ to account for 19,052 genes tested in the gene-based analyses.

### Polygenic score calculations

In addition to the primary GWAS analyses described above, we also calculated polygenic scores (PGS) from UK Biobank GWAS results of four cancer-related phenotypes relevant in the context of our study and assessed how much of the variance of the MMC and IR phenotypes can be explained by these PGS. To this end, we obtained summary statistics from the Pan-UKB project v2 (https://pan.ukbb.broadinstitute.org; [52]) of the UK Biobank for several neoplasms analyzed in that study, i.e., “cancer of bronchus”, “malignant neoplasm of female breast”, “breast cancer phenotypes”, and “chemotherapy”. PGS calculations were performed using default parameters in PRSice-2 software [53]. Statistical analyses fitted general linear regression models with PGS as predictor adjusting for principal components 1-3 (PC1-3) as covariates analogous to the main GWAS analyses. Statistical significances for these predominantly exploratory analyses were set to 0.05.

## Results

### Cellular sensitivity towards mitomycin C and ionizing radiation

We examined a total of 140 LCLs derived from older men from a pool of 432 LCLs available from the BASE-II cohort for their sensitivity to MMC and IR. Sensitivity towards MMC and IR was determined in in two independent batches each by two investigators at different timepoints. Each batch comprised 70 cell lines, with the exception of batch 2 for IR for which sensitivity data was generated for 67 cell lines due to insufficient growth of three cell lines. While the overall AUC values of the two batches were approximately normally distributed (Supplementary Figure 1), there were statistically significant differences regarding the phenotype distribution of the two batches (Figure 2), which might be explained most likely by slight differences in dose rates between the two baches that were examined at different times approximately 1.5 years apart. Thus, the two batches were analyzed separately by GWAS, followed by a fixed-effect meta-analysis of the results across the two experimental batches.

**Figure 2:**
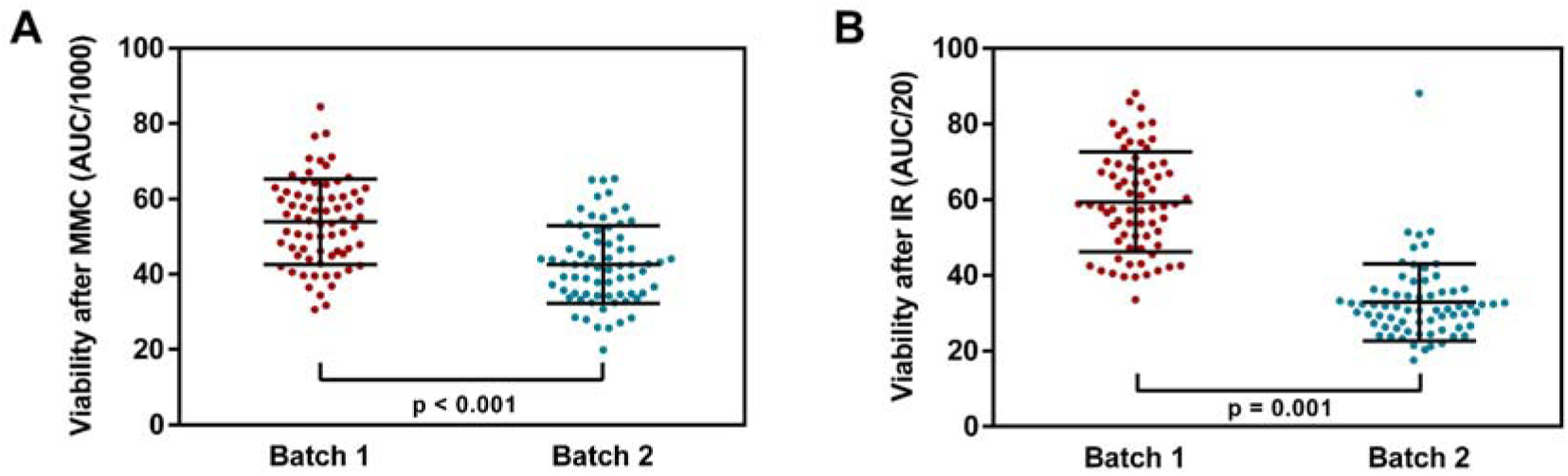
Group differences regarding the phenotype distribution. The column scatter graphs show statistically significant group differences in the phenotype distribution between the two batches. which were examined at different timepoints approximately 1.5 years apart by different examiners (p ≤ 0.001). Viability after MMC treatment (A) and after IR treatment (B) is shown. For illustration purposes, once again AUC/1000 was used to show the viability after MMC treatment and AUC/20 for IR. The correlation between the AUC values of the viability curves after MMC treatment with those after treatment with IR was r = 0.729 (p < 0.001).

### GWAS reveals genome-wide significant association between cellular viability after treatment with ionizing radiation and a variant in *CDH13*

#### SNP-based GWAS analyses

The lambda value for the GWAS meta-analysis of this analysis arm was 1.0486 (Supplementary Figure 2, and 1.0333 and 0.9709 for batch 1 and 2, respectively), suggesting no evidence for a substantial inflation of the genome-wide test statistics. Overall, there were several signals showing statistically suggestive (i.e., p < 1 × 10^-5^) evidence of association, e.g., on chromosomes 1, 3, and 7 (Figure 3A, Supplementary Tables 1-3). Furthermore, one variant, i.e., rs74728080 located in *CDH13* showed genome-wide significant (p < 5 × 10^-8^) association with cellular viability after cell treatment with IR (Figure 3A, Supplementary Table 3). The association was present at sub-genome-wide levels in both experimental batches with effect sizes pointing in the same direction, leading to significant results in the meta-analysis of the results across the entire cohort (p = 3.531 × 10^-8^; ß = 2.81; Figure 3A). The consistency in results across both batches supports the validity of this finding.

**Figure 3:**
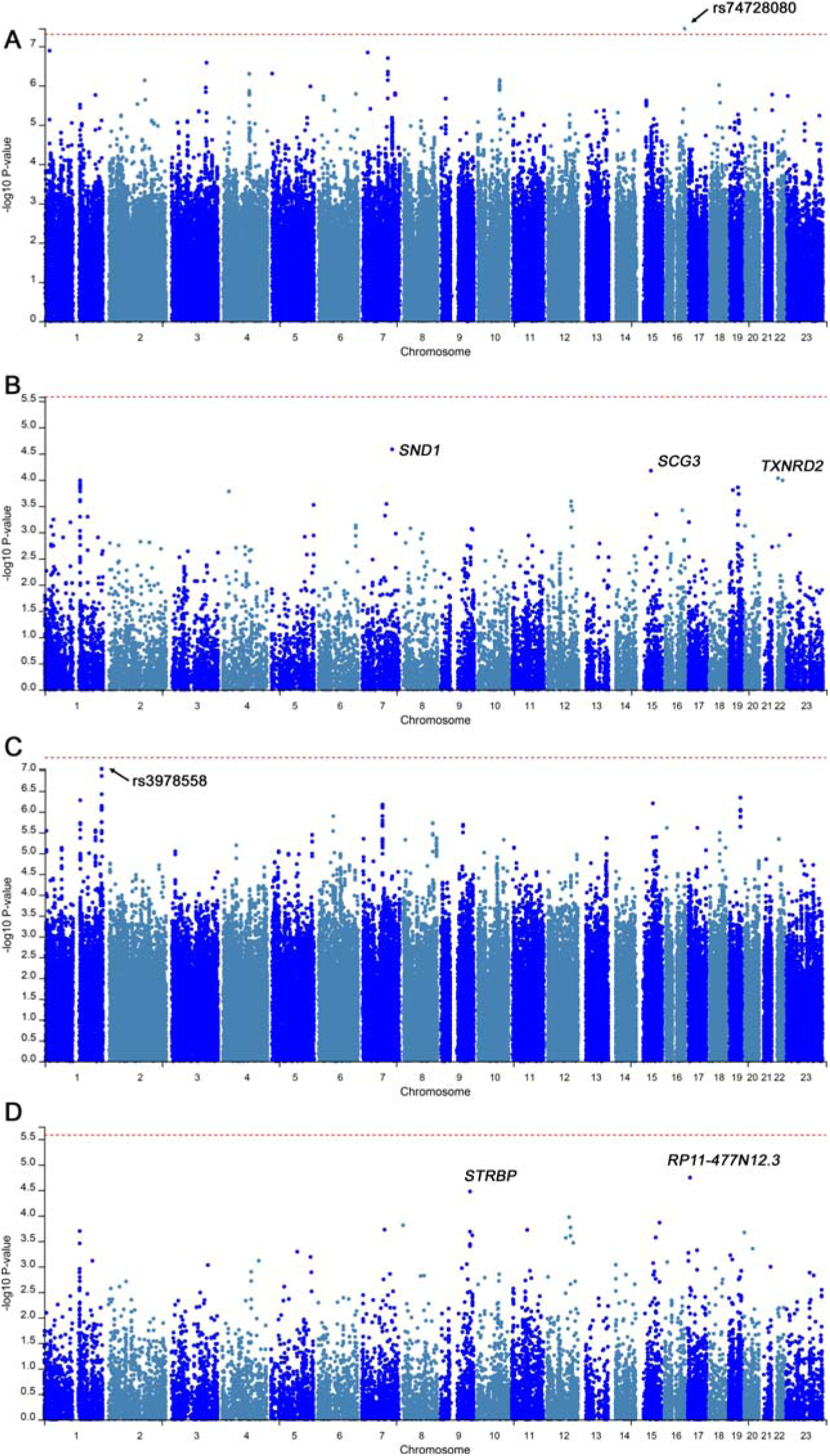
SNP and gene-based meta-analysis on sensitivity towards IR and MMC. Manhattan plots for GWAS meta-analyses (batches 1 and 2) on IR sensitivity (A) SNP-based, (B) gene-based and MMC sensitivity (C) SNP based and (D) gene-based. SNPs or genes are plotted on the x-axis according to their chromosomal location. P-values of 5 × 10^-8^ are marked by the red line. The top associated SNPs and genes are indicated. SNP rs74728080 crosses the threshold of p < 5 × 10^-8^ indicating genome-wide statistical significance for its association with IR sensitivity (A).

According to the Ensembl database (https://www.ensembl.org/index.html; [54]), rs74728080 is located at position 16:83277563 on chromosome 16 (cytogenetic position 16q23.3) in an intron of the *CDH13* (alias *CDHH*; [55]) gene encoding cadherin 13 (CDH13; alias: T-cadherin, H-cadherin (heart); [55]). CDH13 is an atypical member of the cadherin superfamily that is anchored to the surface of the cell membrane and that has no transmembrane or cytoplastic domain [56]. According to dbSNP (https://www.ncbi.nlm.nih.gov/snp/rs74728080), the reference allele for rs74728080 is an adenine (A) base which is changed to a guanine (G) at the variant site [54]. With an MAF of ∼1.3% in non-Finnish Europeans, this SNP is comparatively rare (according to the gnomAD database v4.1.0) [57]. According to the Variant Effect Predictor (VEP) algorithm implemented in Ensembl, the variant allele elicits no known or predicted biological/functional consequences [54]. Similarly, its score on the Combined Annotation Dependent Depletion database (CADD; https://cadd.gs.washington.edu (v1.7); [58]) is comparatively low (PHRED = 0.136), again suggesting no major “biological deleteriousness” of this variant. Owing to its low frequency, no expression quantitative trait loci (eQTL) data are available for rs74728080 on the Genotype Tissue Expression database (GTEx; https://www.gtexportal.org/home/; [59]). Lastly, while the GWAS Catalog (https://www.ebi.ac.uk/gwas/home; [60]), a comprehensive database for GWAS results published in humans, lists a large number of traits showing association with *CDH13*, there are no entries for the lead variant highlighted in our GWAS.

#### Gene-based GWAS analyses

Unlike the single-variant analyses, the gene-based screening did not reveal any findings showing genome-wide significance (p < 2.62×10^-6^; Figure 3B, Supplementary Table 4). The strongest signals in this arm of our project were observed with variants in *SND1* (p = 2.62×10^-5^), *SCG3* (p = 6.70×10^-5^), and *TXNRD2* (p=9.31×10^-5^) (Table S1).

According to the HUGO Gene Nomenclature Committee (HGNC; [55]), *SND1* (alias: *TDRD11*, *p100*) is a protein-coding gene on chromosome 7q32.1. The gene product is staphylococcal nuclease and tudor domain containing 1 (SND1), also known as Tudor-SN or p100 Epstein-Barr virus nuclear antigen 2 (EBNA2) co-activator [55]. The protein serves as a transcriptional co-activator [61] and interacts with other transcription factors, including STAT5 and STAT6 [62–64]. The GWAS Catalog [60] lists nearly 50 traits that show genome-wide significant association with variants in this gene, mostly from neuropsychiatric fields, but none related to oncology/neoplasms.

Second, *SCG3* (alias: *SgIII*) is located on chromosome 15q21.2 and encodes secretogranin III (SCG3; [55]). The protein belongs to the chromogranin/secretogranin family of neuroendocrine secretory proteins. SCG3 plays a role in peptide hormone-producing cells and is involved in the formation of secretory granules. However, its function has not yet been fully understood [65]. The GWAS Catalog [60] lists 15 traits associated with variants in this gene, however, presumably none with greater relevance here. Third, *TXNRD2* encodes thioredoxin reductase 2 (alias: *TR*, *TRXR2*, *TR3*, *SELZ*, *TXNR2*), is located on chromosome 22q11.21 [55], and represents a selenoprotein involved in regulation of intracellular redox homeostasis and free radical scavenging [66]. Additionally, TXNRD2 is attributed an important role in heart function [67, 68]. The GWAS Catalog lists mostly traits related to eye diseases such as glaucoma from 30 studies [60].

Lastly, while the intronic variant rs74728080 within *CDH13* reached genome-wide significance in single-SNP analyses, the gene-based test performed in this arm of our study showed only moderate evidence for association (P=0.171). This discrepancy is not unexpected, as the signal from a single low-frequency variant is prone to dilution by the large number of neutral (i.e. non-associated) markers included in mean-based gene aggregation analyses that are implemented in MAGMA.

### GWAS reveals several sub-genome-wide significant association signals with cellular viability after mitomycin C treatment

#### SNP-based GWAS analyses

Similar to the IR GWAS analyses, the genome-wide lambda values of the MMC GWAS do not suggest an inflation of the test statistics (1.0414 for the meta-analysis (Supplementary Figure 2), 1.0059 for batch 1, and 1.0086 for batch 2). The most interesting signal was elicited by SNP rs113978558 in an intron of the *PLD5* gene at position 1:242098270 on the forward strand of chromosome 1 (cytogenetic position 1q43). The reference allele of this SNP is a thymine (T) at the given location, which is changed to a guanine (G) as the alternative allele [54]. The variant missed the genome-wide significance threshold by only a small margin (p = 9.232 × 10^-8^; ß = 1.44; Figure 3C). The MAF is ∼0.05 in the non-Finnish European population according to the gnomAD database [57] and ∼0.04 in our dataset. In addition, there were several additional genome-wide suggestive signals in this GWAS with markers located on chromosomes 1, 7, 15, and 19 (Figure 3C, Supplementary Tables 5-7).

*PLD5* encodes phospholipase D family member 5 (PLD5). Phospholipase D (PLD) facilitates the hydrolysis of phospholipids to form phosphatidic acid, commonly acting as a secondary messenger in the transduction of intracellular signals [69]. SNP rs113978558 has no major known or predicted impact on protein function according to the VEP [54] or CADD [58] databases. On GTEx v10 [59], this SNP is associated with the expression of ENSG00000272865 in the lung (p=8.2×10^-6^), a novel transcript probably representing an anti-sense long non-coding RNA (lncRNA) within the *PLD5* open reading frame according to the Ensembl database (https://www.ensembl.org/). In contrast, no eQTL or splicing quantitative trait loci (sQTL) associations with this SNP are reported for *PLD5* itself. Finally, while the GWAS Catalog [60] does not contain any entries for the sentinel SNP, the *PLD5* gene itself has been found associated with several human traits, including two related to neoplasms, i.e., “family history of lung cancer” and “acute myeloid leukemia”, tentatively suggesting a potential role in cancer development.

#### Gene-based GWAS analyses

In this arm of our study, there are two noteworthy signals that emerged in the gene-based analyses, i.e., with *RP11-477N12.3* (p = 1.80×10^-5^) and *STRBP* (p = 3.36×10^-5^). Despite the comparatively strong overall associations, we note that these miss the threshold of genome-wide significance (p < 2.62 × 10^-6^; Figure 3D, Supplementary Table 8) by about one order of magnitude. According to the Ensembl database, *RP11-477N12.3* is annotated to transcript ENST00000399363 which encodes germ cell-specific gene 1-like protein 2. The HGNC-approved symbol is *GSG1L2* and the gene is located on chromosome 17p13.1 [55]. It is predicted to be an integral component of the plasma membrane [70]. However, to the best of our knowledge, no known disease-related biological function has been attributed to this gene. Interestingly, the GWAS Catalog [60] lists “asparaginase hypersensitivity in acute lymphoblastic leukemia” as one of only four traits associated with the gene locus, indicating a possible role in cancer development.

The second gene-based signal was elicited by variants in *STRBP* (alias *SPNR*; previously *ILF3L*) encoding for spermatid perinuclear RNA binding protein (STRBP), also known as interleukin enhancer binding factor 3-like (ILF3L). The gene is located on chromosome 9q33.1-q33.3 [55]. As the name suggests, the protein is involved in spermatogenesis and sperm function, particularly in the regulation of cell growth [71]. Schumacher et al. suggest that the protein enables RNA-binding activity by acting as a link between stored RNAs and microtubules that mediate their transport and final translation [72]. The GWAS catalog lists several traits associated with variants in *STRBP*, most of these associated with height and BMI, but also some cancer-related traits such as astrocytoma and glioma [60].

#### PGS calculations from GWAS of four cancer-related traits in Pan-UKB explain a significant fraction of phenotypic variance of IR- and MMC-induced cell viability

For cell viability after IR treatment, the PGS calculations showed nominally significant associations with three out of the four traits selected from the Pan-UKB project (i.e., all but “chemotherapy”). The strongest association was observed with the AUC of the IR viability curve and “cancer of bronchus” (R^2^ ∼4.9%, p ∼0.01; Supplementary Table 9, Figure 4). Interestingly, the PGS analyses for cell viability after MMC treatment only showed nominally significant associations with the trait “chemotherapy” (R^2^ ∼3%, p ∼0.03), but none of the other primary cancer traits (Supplementary Table 10, Figure 4). Collectively, these data show a moderate, but nominally significant correspondence between genetic effects underlying various cancers and cancer-related traits in the Pan-UKB project [52] and our *in vitro* experiments, predominantly those related to the IR treatment.

**Figure 4:**
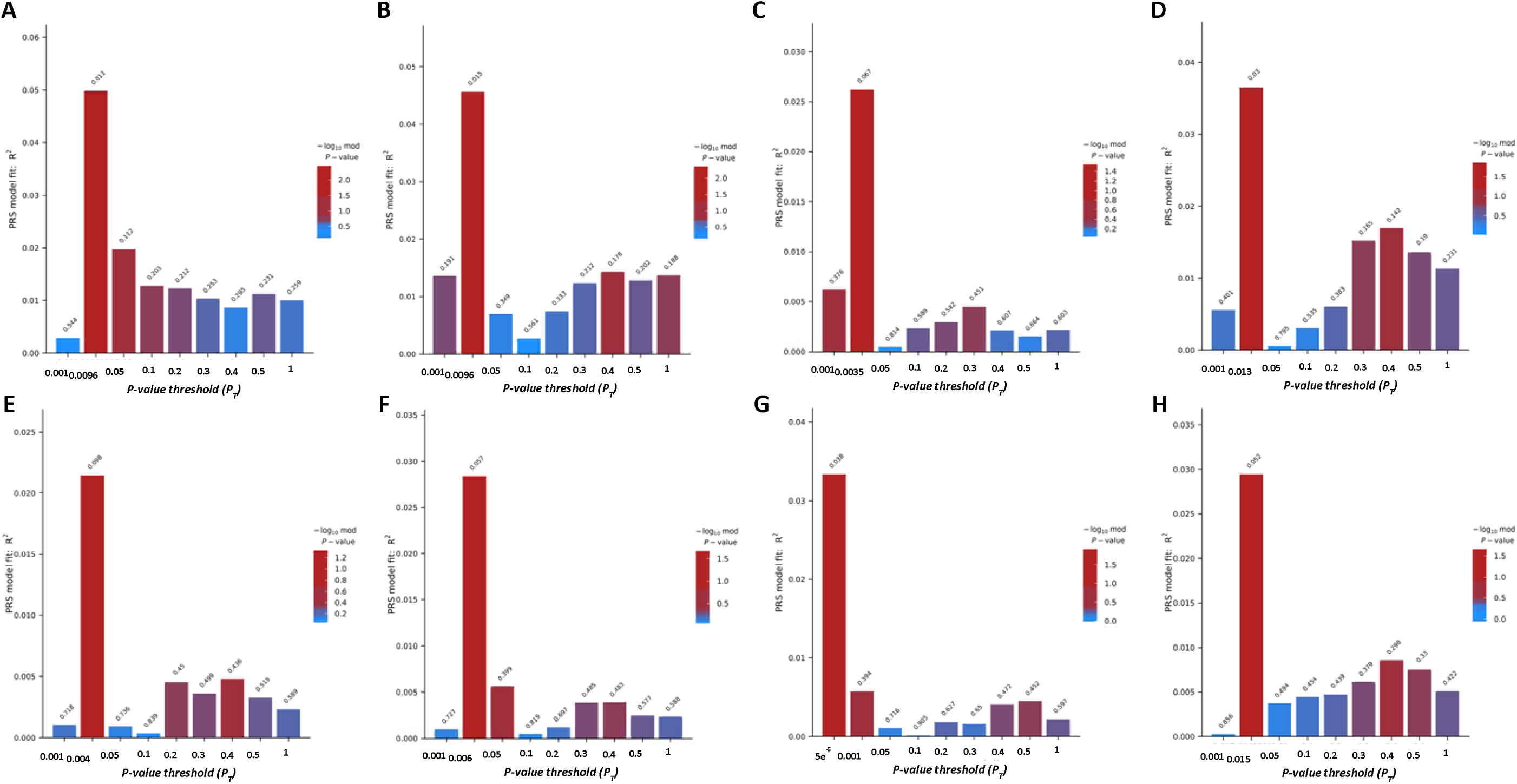
PGS calculated from GWAS of four cancer-related traits in Pan-UKB explain a significant fraction of phenotypic variance of IR- and MMC-induced cell viability. PGS calculation for IR-AUC (A-D) and MMC-AUC (E-H) using summary statistics from the trait (A, E) Cancer of bronchus; lung (UKBB); (B, F) Malignant neoplasm of female breast (UKBB); (C, G) Chemotherapy (UKBB); (D, H) Breast cancer (UKBB).

## Discussion

In the current study, we assessed the viability of LCLs after treatment with MMC and IR and searched for genetic determinants of this outcome by GWAS. For IR treatment, the GWAS analyses identified a comparatively rare variant, rs74728080, in *CDH13* that was significantly associated with cellular radiosensitivity at the genome-wide level, suggesting a protective effect of the variant against IR-induced cellular damage. For MMC treatment, our GWAS analyses showed strong association with markers in the *PLD5* gene, although these did not pass the genome-wide significance threshold.

The gene product of *CDH13* is cadherin 13, an atypical member of the cadherin superfamily. Different from most other cadherins, it misses the transmembrane and intracellular domains, is bound to the plasma membrane by a glycosylphosphatidylinositol (GPI) anchor and is not enriched in adherens junctions [73]. Philippova et al. showed that CDH13 is not essential for the maintenance of intercellular adhesion but is thought to function primarily as a signaling molecule [74]. The protein protects vascular endothelial cells from oxidative stress-induced apoptosis and is associated with resistance to atherosclerosis (reviewed in [26]). Joshi et al. investigated the functional relationship between oxidative stress, *CDH13* expression, and cell survival status using cultures of human umbilical vein endothelial cells (HUVEC) [75]. CDH13 levels were shown to be upregulated due to oxidative stress. Their results suggest that overexpression leads to increased cell survival, which is due to activation of the PI3K/Akt/mTOR survival signaling pathway and simultaneous suppression of the proapoptotic p38 MAPK pathway [75]. IR, as used in the current study, is well-known to induce oxidative stress in exposed cells (e.g., [76]). Additionally, *CDH13’s* role as tumor suppressor is reduced by hypermethylation, a molecular read-out observed in many cancers (e.g., [27–33]), and has also been described as a risk factor for neurological and psychiatric disorders, among them attention-deficit/hyperactivity disorder (ADHD) and autism spectrum disorders, as well as major depression (reviewed, e.g., in [77]). Furthermore, CDH13 functions as a negative regulator of axon growth during neural differentiation [78, 79]. Lastly, while the lead SNP of the *CDH13* association identified here (i.e. rs74728080) is not included in the GWAS catalog, it is listed as a genetic determinant of “skin changes due to chronic exposure to non-ionising radiation” (p = 2.28 × 10^-6^) in the PhenoScanner database (http://www.phenoscanner.medschl.cam.ac.uk/) based on GWAS results in the UK Biobank dataset. While this finding was specifically made with non-ionizing radiation, i.e. electromagnetic radiation without enough energy to ionize atoms or molecules, it may point to a more general radiation-modifying effect of the CDH13 protein, in line with the findings of our study.

Despite the strong statistical support of the association between rs74728080 and cell viability after IR treatment, we note that this is a “singleton” association signal, i.e., there are no other correlated variants in the *CDH13* region, which typically characterize *bona fide* GWAS results. While this is likely due to the fact that rs74728080 is relatively rare in this dataset (MAF = 0.013), in agreement with frequency data in the European population (MAF = 0.01283; https://gnomad.broadinstitute.org; [57]), this relative infrequency also increases the likelihood of a false-positive finding.

In summary, although rs74728080 meets the formal criteria of genome-wide significant association in the context of our analyses, these findings need to be replicated in independent datasets. Notwithstanding, *CDH13* is a plausible candidate in the context of our study due to its protective role in oxidative stress-induced apoptosis (reviewed in [26]) and carcinogenesis (e.g., [27–33]).

The second most interesting finding of our GWAS relates to a variant (i.e. rs113978558) in the *PLD5* gene which was associated with decreased cellular sensitivity to MMC. However, as stated above, the statistical support of this finding did not formally reach genome-wide significance and, accordingly, must be interpreted with caution and require replication in an independent dataset.

*PLD5* belongs to the family of phospholipase D enzymes which catalyze the hydrolysis of phosphodiester bonds in glycerophospholipids and have been reported as critical regulators of cell proliferation, survival signaling, cell transformation, and tumor progression (e.g., reviewed in [80]). More specifically, PLD5 has been described in the context of cell proliferation and metastasis in prostate cancer [81], pro-inflammatory effects in chronic obstructive pulmonary disease (COPD) [82], uterine fibroids [83, 84], autism spectrum disorder [85], immune thrombocytopenia (ITP) [86], or corpus callosum malformations [87]. Liu et al. reported evidence for downregulation of microRNA miR-145-5p in prostate cancer cell lines and tissues and modulation of PLD5 oncogenic effects mediated by this microRNA [81].

In summary, *PLD5* appears as a reasonable candidate to be functionally involved in cell viability, although the genetic evidence generated in our study must be interpreted with caution until independent replication is provided. Unlike the *CDH13* signal in our IR GWAS, the *PLD5* variant left a “trail” of highly significant association results correlated with the sentinel variant – likely owing to its higher MAF – which reduces the overall likelihood of this signal representing a false-positive finding. At this time, we can only speculate about the functional mechanism underlying this association. According to some studies (e.g., [38]), an overexpression might promote carcinogenesis due to downregulated apoptosis, which might also be the predominant mechanism in the context of our MMC-based cell viability assay.

In contrast to the SNP-based results, the gene-based GWAS analyses did not reveal any genome-wide significant signals and are therefore not highlighted in this discussion section.

In the last analysis arm of our study, we found that PGSs constructed from previous, cancer-related GWAS analyses in the Pan-UKB project [52] show moderate, but statistically significant concordance to our *in vitro* experiments. In particular for the IR-based cell viability assays, the PGS analyses suggest that genetic variants predisposing to lung and breast cancer also significantly explain a noteworthy portion of phenotypic variance (∼3.5-5%) of our experimental cancer-related phenotypes. In general, these results support the main conclusions of our study using a different methodology and genomic data from an independent dataset.

A major strength of our study is that LCL sensitivity was assessed in triplicate within each experiment and across three independent experiments for each cell line and treatment (MMC and IR), thereby ensuring high-quality data and minimizing the effects of experimental and biological variation. We also analyzed the two batches separately by GWAS, followed by a fixed-effect meta-analysis of the results across the two experimental batches. Despite the strength of the identified associations and the good correspondence with related GWAS findings from the UK Biobank, our study is subject to a number of limitations. First and foremost, we need to highlight the exceedingly small sample size of our GWAS. Small sample sizes reduce the power to detect true genetic effects (type-II error), but also increase the likelihood of false-positive findings (type-I error). Despite this limitation, we note that our outcome phenotype was the result of a comparatively laborious experimental paradigm for which it would have been prohibitive to generate more data. Eventually – and this is also true for GWAS with much larger sample sizes – only replication in comparable independent datasets will show which of the results presented here will prove to be genuine. Second, to obtain a homogeneous cohort and, thus, stand the highest chance of identifying genetic associations, we performed the viability assays using cell cultures only from a selected subgroup, namely LCLs from older male BASE-II participants. Therefore, it remains uncertain to what extent the results can be transferred to women and/or other cell types. Lastly, we emphasize that associations identified in a GWAS design are statistical and allow no inference on causality or functional mechanisms. Thus, in addition to the independent replication of the primary association results, our findings will need to be assessed in dedicated functional experiments (e.g., using siRNA-based knockdown) to delineate molecular mechanisms.

## Conclusion

In conclusion, our GWAS analyses of cell viability suggest that one (or more) variants in *CDH13* may lower cellular sensitivity to IR-induced cellular stress. Based on previous studies, we speculate that the associated variant mediates higher *CDH13* expression, resulting in higher resistance to IR-induced oxidative stress. Future work should focus on independent replication of these promising findings and on elucidating the precise biological role of CDH13 in cellular sensitivity to IR, tumorigenesis, and resistance to therapeutic mutagens.

## Conflict of interest

The authors declare no competing interests.

## Funding

This work was supported by a grant of the Deutsche Forschungsgemeinschaft to ID (grant number DE 842/4-1). This article uses data and cell lines from the Berlin Aging Study II (BASE-II). BASE-II was supported by the German Federal Ministry of Education and Research under grant numbers #01UW0808; #16SV5536K, #16SV5537, #16SV5538, #16SV5837, #01GL1716A, and #01GL1716B.

## Authors’ contributions

Conceptualization: H-L.S., LB, I.D.; Data curation: L.B., I.D.; Formal analysis: H.-L.S., O.O.; Investigation: H.-L.S., O.O.; Methodology: S.H., B.S., L.B., I.D.; Project administration: I.D.; Resources: S.H., B.S., L.B., I.D.; Supervision: L.B., I.D.; Visualization: H.-L.S., O.O., I.D.; Writing - original draft: H.-L.S., L.B, I.D.; Writing - review & editing: all authors.

## Data Availability

Due to participant privacy concerns, BASE-II data are available only upon reasonable request. Additional information can be found on the BASE-II website: https://www.base2.mpg.de/7549/data-documentation.

## Notes

### Competing Interest Statement

The authors have declared no competing interest.

### Author Declarations

The Ethics Committee of the Charite - Universitaetsmedizin Berlin approved the study (approval numbers EA2/029/09 and EA2/144/16).

