## Supplementary Figures for "*CDH13* is associated with cellular viability after exposure to ionizing radiation using genome-wide screening"

**Supplementary Figure 1**

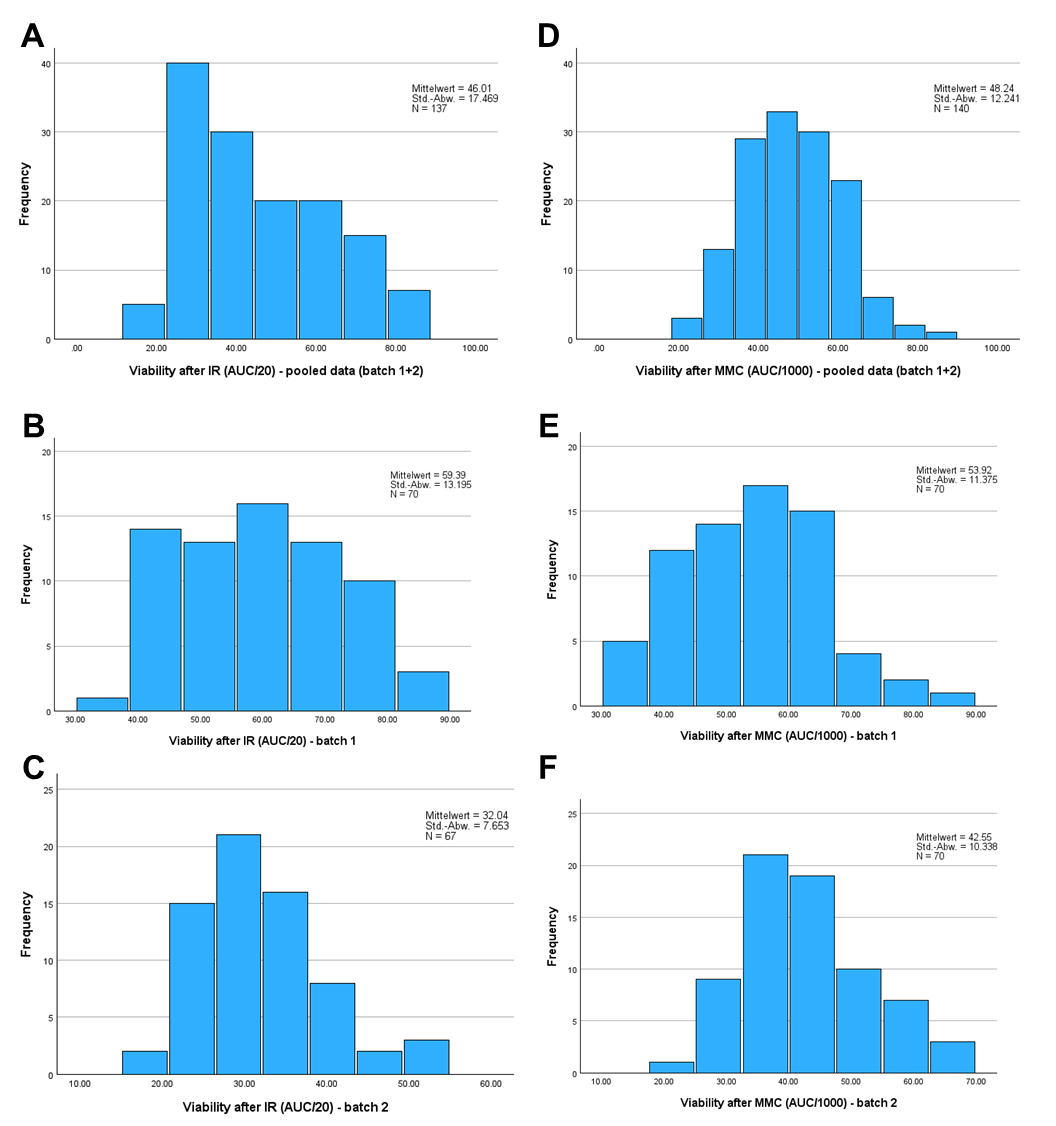

**Supplementary Figure 1: Phenotype distribution**

(A) through (C) show the viability after IR treatment, whereas (D) through (F) show the viability after MMC treatment. More precisely, the x-axes show the viability after MMC/IR treatment as indicated by the AUC values of the viability curves of the cell lines. For illustration purposes, AUC/20 was used to show the viability after IR treatment and AUC/1000 for MMC. The y-axes show the respective frequencies. (A) and (D) respectively show the values for the pooled data (batch 1 + 2), (B) and (E) only for batch 1 and (C) and (F) only for the batch 2. The figure also includes the sample sizes as well as the mean values and standard deviations (SD).

**Supplementary Figure 2**

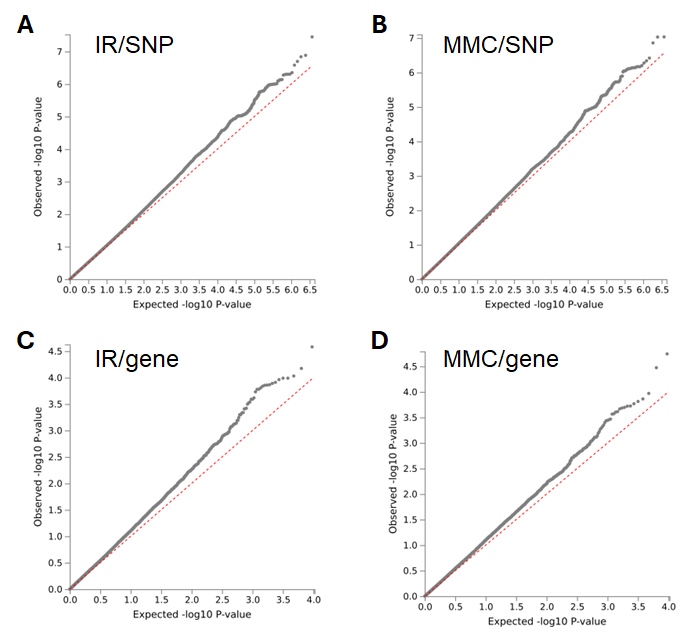

**Supplementary Figure 2: Quantile-quantile plots from the GWAS of IR-AUCs and MMC-AUCs for SNP the SNP-based and gene-based meta-analyses**
